# Renin, Major Adverse Cardiovascular Outcomes, and the Use of Mineralocorticoid Receptor Antagonists in Hypertension

**DOI:** 10.64898/2026.05.11.26352940

**Authors:** Cheng-Hsuan Tsai, Yu-Ching Chang, Chin-Chen Chang, Andrew J Newman, Jenifer M Brown, Vin-Cent Wu, Yen-Hung Lin, Anand Vaidya

## Abstract

**Background and Aims:** The prognostic significance of renin activity and its role in identifying patients who may benefit from mineralocorticoid receptor antagonists (MRAs) remain incompletely defined.

**Methods:** In a multicentre retrospective cohort of 16,600 adults with hypertension, we evaluated associations between baseline plasma renin activity and major adverse cardiovascular events (MACE), comprising ischaemic stroke, intracranial haemorrhage, myocardial infarction, and all-cause death. Renin was analysed using restricted cubic splines and categories. Time-dependent Cox models assessed associations between MRA use and outcomes across renin strata, treating MRA exposure as time-varying.

**Results:** Renin showed a U-shaped association with MACE, with the lowest estimated risk at 1.17 ng/mL/h. Compared with renin 0.3–1.0 ng/mL/h, both very low renin (<0.3 ng/mL/h; adjusted hazard ratio [HR] 1.29, 95% CI 1.15–1.45) and high renin (>3.0 ng/mL/h; HR 1.19, 1.06–1.33) were associated with higher MACE risk. MRA use was associated with lower MACE risk when renin was very low (<0.3 ng/mL/h; HR 0.59, 95% CI 0.46–0.74) or low (0.3–1.0 ng/mL/h; HR 0.71, 95% CI 0.55–0.92), whereas no significant association was observed when renin was high (>3.0 ng/mL/h; HR 1.07, 95% CI 0.76–1.52). MRA use was associated with lower all-cause mortality across renin categories ≤3.0 ng/mL/h, whereas no significant association was observed at renin >3.0 ng/mL/h.

**Conclusion:** Baseline renin showed a U-shaped association with MACE. The use of MRAs was associated with lower risk of MACE when renin was low and very low. These findings provide support for renin-guided MRA therapy in hypertension.

## Introduction

Hypertension is the leading modifiable risk factor for cardiovascular morbidity and mortality worldwide^1,2^. However, optimal risk stratification and treatment selection remain major clinical challenges. Despite the availability of multiple antihypertensive therapies, substantial inter-individual variability in treatment response and persistently suboptimal blood pressure control underscore the need for more precise, mechanism-based approaches to hypertension management^3–5^.

Plasma renin has long been recognized as a central biomarker reflecting the physiological state of the renin-angiotensin-aldosterone system (RAAS)^6^. In clinical practice, renin measurement plays an established role in screening for primary aldosteronism (PA), a condition characterized by low-renin hypertension and renin-independent aldosterone production that drives sodium retention, volume expansion, and adverse cardiometabolic outcomes^7,8^. Contemporary guidelines recommend broader screening for PA using renin and aldosterone measurements in nearly all people with hypertension. Further, recent guidelines recommend treating PA, a state of low-renin hypertension, with mineralocorticoid receptor antagonists (MRAs) such that renin increases, thereby advising a renin-guided therapeutic approach. Adoption of these guidelines will mean that nearly all of the 1.3 billion people with hypertension will have measurements of renin, highlighting the need to clarify the clinical significance of renin measurements in people with general hypertension, beyond PA alone^1,9,10^.

Accumulating evidence suggests that renin provides broader pathophysiological insights beyond diagnostic classification alone^5,8,11–15^. Low renin has been associated with differential blood pressure responses to antihypertensive therapy, particularly greater responsiveness to MRAs^15,16^. These observations suggest that a low-renin phenotype may identify patients with clinically relevant mineralocorticoid receptor activation, either because it represents unrecognized PA^17^, or a differential state of MR over-activation ^16,18^. Prior clinical and mechanistic studies further support associations of low-renin hypertension with cardiovascular risk and responsiveness to MRA therapy^4,6,13,17,19–28^. Nevertheless, the prognostic significance of renin across its continuous range in general hypertension, and the association between MRA use and long-term clinical outcomes at different baseline renin levels remain incompletely defined.

In this study, we hypothesized that baseline renin is associated with long-term cardiovascular risk and that the association between MRA use and clinical outcomes varies according to baseline renin levels. To examine this hypothesis, we conducted a large multicenter retrospective cohort study. We first characterized the association between plasma renin activity and major adverse cardiovascular events, and then evaluated associations between MRA use during follow-up and clinical outcomes across renin-defined categories using time-varying analyses.

## Methods

### Data source

This retrospective cohort study used data from a multicenter electronic medical record database comprising eight hospitals within National Taiwan University Hospital (NTUH) healthcare system across Taiwan, including both medical centers and regional hospitals. These hospitals represent diverse healthcare settings across different geographic regions and levels of urbanization in Taiwan, enhancing the generalizability of the study findings. The database is maintained and curated by NTUH with standardized data management procedures to ensure data accuracy and completeness. Disease diagnoses were identified using the International Classification of Diseases, Ninth Revision, Clinical Modification (ICD-9-CM) codes prior to 2015, and transitioned to the Tenth Revision (ICD-10-CM) starting in 2016 (**Supplementary Table 1**). The study was approved by the Research Ethics Committee of National Taiwan University Hospital, Taipei, Taiwan (approval number: 202504119RINB). The requirement for informed consent was waived due to the retrospective nature of the study and the use of de-identified patient data.

### Study Population

The data for this study encompassed the medical records of hypertensive patients who underwent plasma renin activity testing at NTUHs. We identified all adult patients with hypertension who underwent plasma renin activity testing between January 1, 2006, and December 31, 2022. We excluded the first and last calendar years from cohort inclusion to ensure at least a 1-year window for baseline covariate assessment and outcome ascertainment. Hypertension was defined by a combination of ICD codes and the documented use of one or more antihypertensive agents. We excluded patients with prior adrenalectomy, baseline use of MRAs, baseline use of loop diuretics, as an indicator of severe heart failure, or missing aldosterone measurements. After applying these criteria, a total of 16,600 patients were included in the final analysis (**Supplementary Figure 1**).

### Study design and analytical framework

We conducted two complementary analyses within a common source cohort to address distinct questions: the association between baseline plasma renin activity and subsequent clinical outcomes, and the association between time-varying MRA use and outcomes within baseline renin categories (**Figure 1**). For both analyses, the index date was the date of baseline plasma renin activity measurement.

**Figure 1.**
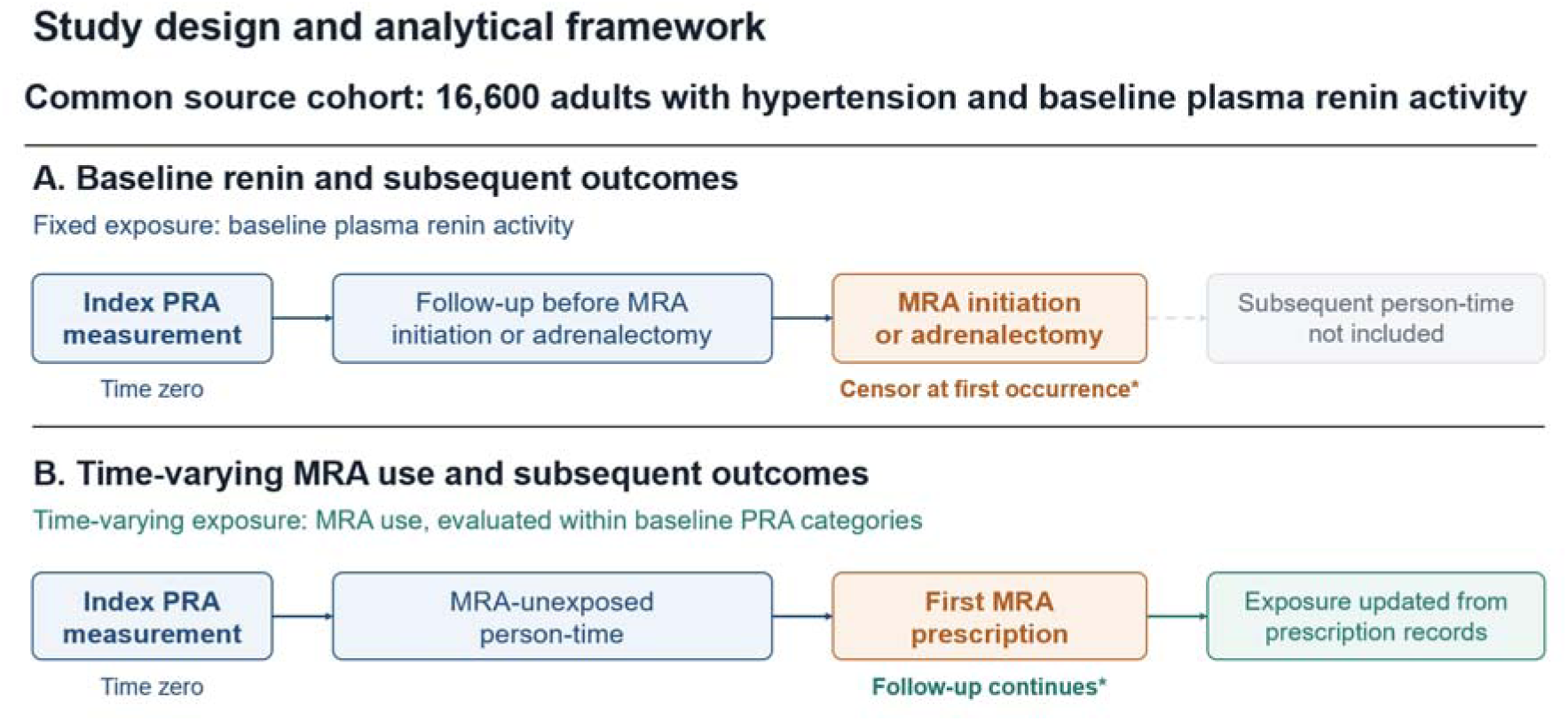
Study design and analytical framework. Both analyses included 16,600 adults with hypertension, with the index PRA measurement defining time zero. (A) Baseline PRA was evaluated using Cox models with restricted cubic splines and categories, adjusting for baseline covariates. Follow-up was censored at MRA initiation or adrenalectomy. (B) Time-varying MRA use was evaluated within baseline PRA categories using time-dependent Cox models adjusted for baseline age and sex and time-varying clinical covariates. The primary analysis censored follow-up at adrenalectomy; the sensitivity analysis did not. *In both analyses, follow-up could end earlier at the outcome of interest, loss to follow-up, or December 31, 2023. MRA, mineralocorticoid receptor antagonist; PRA, plasma renin activity.

In the baseline renin analysis, plasma renin activity was treated as a fixed exposure and evaluated continuously and categorically. Follow-up was censored at the first initiation of MRA therapy or adrenalectomy, allowing assessment of outcomes occurring before initiation of these treatments.

In the MRA analysis, follow-up was divided into consecutive 3-month intervals. MRA exposure was treated as a time-varying variable and classified for each interval according to prescription records. Patients could contribute both MRA-exposed and MRA-unexposed intervals during follow-up, with unexposed person-time serving as the reference within each baseline renin category. Follow-up continued after MRA initiation, with exposure status and clinical covariates updated during follow-up. The primary analysis censored patients at adrenalectomy; a sensitivity analysis continued follow-up without censoring at adrenalectomy.

### Covariates and Outcomes

Baseline characteristics were collected for this study, including demographics (e.g., age, sex, body mass index), laboratory parameters (e.g., serum creatinine and uric acid), comorbidities (e.g., diabetes and chronic kidney disease), cardiovascular history (i.e., heart failure), and baseline antihypertensive medication use (detailed in **Table 1**). For the MRA analyses, body mass index, serum creatinine, diabetes mellitus, heart failure, and concomitant antihypertensive medication use were updated during follow-up and included as time-varying covariates. Antihypertensive medications included renin-angiotensin system inhibitors, beta-blockers, alpha-blockers, vasodilators, thiazide diuretics, calcium channel blockers, clonidine, and minoxidil. Age and sex were included as baseline covariates.

**Table 1.** Demographic and clinical characteristics of patients receiving plasma renin activity examination.

|  | Total | Renin <0.3 | Renin 0.3–1.0 | Renin >1.0–3.0 | Renin >3.0 |
|---|---|---|---|---|---|
| Variable | (*n* = 16,600) | (*n* = 3,617) | (*n* = 3,748) | (*n* = 5,464) | (*n* = 3,771) |
|---|---|---|---|---|---|
| Age, year | 54.2 ± 16.1 | 58.3 ± 14.5 | 56.6 ± 15.0 | 52.9 ± 16.4 | 50.1 ± 16.9 |
| Age >65 years | 4,487 (27.0) | 1,189 (32.9) | 1,136 (30.3) | 1,383 (25.3) | 779 (20.7) |
| Male sex | 8,529 (51.4) | 1,620 (44.8) | 1,792 (47.8) | 2,903 (53.1) | 2,214 (58.7) |
| Body weight, kg | 68.9 ± 15.6 | 66.5 ± 14.7 | 68.1 ± 15.2 | 69.8 ± 15.8 | 70.9 ± 16.4 |
| Body height, cm | 163.3 ± 9.0 | 162.0 ± 8.7 | 162.6 ± 9.0 | 163.8 ± 9.0 | 164.4 ± 9.1 |
| Body mass index, kg/m<sup>2</sup> | 25.7 ± 4.6 | 25.2 ± 4.3 | 25.6 ± 4.5 | 25.8 ± 4.6 | 26.0 ± 4.8 |
| Biochemical studies |  |  |  |  |  |
| Plasma renin activity, ng/mL/h | 1.2 [0.4, 2.6] | 0.13 [0.08, 0.20] | 0.60 [0.44, 0.80] | 1.6 [1.3, 1.9] | 5.9 [4.4, 7.1] |
| Plasma aldosterone concentration, ng/dL | 25.5 [15.6, 37.5] | 22.9 [13.2, 35.5] | 23.0 [14.1, 33.4] | 24.8 [16.2, 34.9] | 33.1 [19.5, 51.0] |
| Aldosterone-to-renin ratio, ng/dL per ng/mL/h | 20.3 [9.2, 58.6] | 177.3 [85.8, 462.2] | 37.7 [22.9, 60.9] | 15.5 [10.0, 22.3] | 5.4 [2.7, 9.4] |
| Hemoglobin, g/dL | 13.4 ± 2.2 | 13.1 ± 2.1 | 13.3 ± 2.1 | 13.6 ± 2.2 | 13.7 ± 2.2 |
| Serum creatinine, mg/dL | 1.08 ± 0.99 | 1.04 ± 0.95 | 1.08 ± 1.05 | 1.07 ± 0.95 | 1.14 ± 1.00 |
| eGFR, mL/min/1.73m<sup>2</sup> | 86.4 ± 30.9 | 87.0 ± 30.7 | 85.7 ± 30.2 | 87.0 ± 30.4 | 85.4 ± 32.3 |
| Sodium, mEq/L | 138.9 ± 3.6 | 139.3 ± 3.7 | 139.1 ± 3.6 | 138.9 ± 3.4 | 138.5 ± 3.6 |
| Potassium, mEq/L | 4.1 ± 0.5 | 4.0 ± 0.6 | 4.1 ± 0.5 | 4.1 ± 0.5 | 4.1 ± 0.5 |
| HbA1c, % | 6.2 ± 1.2 | 6.2 ± 1.3 | 6.2 ± 1.2 | 6.2 ± 1.2 | 6.2 ± 1.2 |
| Fasting glucose, mg/dL | 105.9 ± 28.7 | 106.3 ± 28.9 | 105.7 ± 28.2 | 105.8 ± 28.3 | 106.1 ± 29.7 |
| Total cholesterol, mg/dL | 187.4 ± 39.1 | 184.6 ± 38.9 | 187.0 ± 38.4 | 188.5 ± 39.9 | 189.1 ± 39.0 |
| Low-density lipoprotein, mg/dL | 111.4 ± 33.0 | 109.0 ± 32.1 | 110.7 ± 32.2 | 112.1 ± 33.6 | 113.5 ± 33.7 |
| High-density lipoprotein, mg/dL | 48.4 ± 13.2 | 48.2 ± 13.4 | 48.7 ± 13.4 | 48.9 ± 13.3 | 47.6 ± 12.7 |
| Triglyceride, mg/dL | 143.2 ± 87.3 | 138.7 ± 84.3 | 138.3 ± 82.5 | 145.1 ± 88.8 | 149.8 ± 92.3 |
| ALT, U/L | 26.8 ± 22.0 | 25.8 ± 22.0 | 26.1 ± 20.8 | 26.7 ± 21.5 | 28.8 ± 23.8 |
| Uric acid, mg/dL | 6.1 ± 1.7 | 5.8 ± 1.7 | 6.0 ± 1.7 | 6.1 ± 1.7 | 6.4 ± 1.8 |
| Comorbidity |  |  |  |  |  |
| Diabetes mellitus | 2,439 (14.7) | 575 (15.9) | 542 (14.5) | 818 (15.0) | 504 (13.4) |
| Hyperlipidemia | 3,144 (18.9) | 720 (19.9) | 702 (18.7) | 1,061 (19.4) | 661 (17.5) |
| Atrial fibrillation | 240 (1.4) | 69 (1.9) | 50 (1.3) | 82 (1.5) | 39 (1.0) |
| Obstructive sleep apnea | 468 (2.8) | 113 (3.1) | 104 (2.8) | 183 (3.3) | 68 (1.8) |
| Chronic obstructive pulmonary disease | 417 (2.5) | 93 (2.6) | 103 (2.7) | 118 (2.2) | 103 (2.7) |
| Chronic kidney disease | 1,495 (9.0) | 337 (9.3) | 357 (9.5) | 487 (8.9) | 314 (8.3) |
| Cardiovascular history |  |  |  |  |  |
| Heart failure | 188 (1.1) | 48 (1.3) | 38 (1.0) | 50 (0.9) | 52 (1.4) |
| Acute myocardial infarction | 98 (0.6) | 21 (0.6) | 19 (0.5) | 32 (0.6) | 26 (0.7) |
| Coronary artery disease | 1,295 (7.8) | 310 (8.6) | 303 (8.1) | 365 (6.7) | 317 (8.4) |
| Intracranial hemorrhage | 480 (2.9) | 122 (3.4) | 85 (2.3) | 161 (2.9) | 112 (3.0) |
| Ischemic stroke | 873 (5.3) | 237 (6.6) | 212 (5.7) | 238 (4.4) | 186 (4.9) |
| Anti-hypertensive medications at baseline |  |  |  |  |  |
| RAS inhibitor | 8,489 (51.1) | 1,789 (49.5) | 1,853 (49.4) | 2,736 (50.1) | 2,111 (56.0) |
| Beta-blocker | 5,489 (33.1) | 1,355 (37.5) | 1,249 (33.3) | 1,670 (30.6) | 1,215 (32.2) |
| Alpha-blocker | 3,036 (18.3) | 796 (22.0) | 697 (18.6) | 930 (17.0) | 613 (16.3) |
| Vasodilator | 835 (5.0) | 184 (5.1) | 182 (4.9) | 312 (5.7) | 157 (4.2) |
| Thiazide | 1,712 (10.3) | 331 (9.2) | 357 (9.5) | 513 (9.4) | 511 (13.6) |
| Calcium channel blocker | 9,370 (56.4) | 2,128 (58.8) | 2,111 (56.3) | 2,977 (54.5) | 2,154 (57.1) |
| Others (Clonidine, Minoxidil) | 381 (2.3) | 109 (3.0) | 89 (2.4) | 86 (1.6) | 97 (2.6) |
| Number of medication | 1.8 ± 1.4 | 1.9 ± 1.5 | 1.7 ± 1.4 | 1.7 ± 1.4 | 1.8 ± 1.4 |

Abbreviation: ALT, alanine aminotransferase; eGFR, estimated glomerular filtration rate; HbA1c, glycated hemoglobin; RAS, renin-angiotensin system.
Data were presented as frequency (percentage), mean ± standard deviation or median [25<sup>th</sup>, 75<sup>th</sup> percentiles].

The primary outcome was major adverse cardiovascular events (MACE), defined as a composite of ischemic stroke, intracranial hemorrhage, myocardial infarction, and all-cause mortality. All-cause mortality was ascertained through linkage with the National Death Registry. Clinical events were captured via inpatient ICD codes, which have demonstrated high diagnostic accuracy in previous validation studies using insurance claim data^29–31^.

Follow-up began on the index date and continued until the first occurrence of the outcome of interest, an analysis-specific censoring event, loss to follow-up, or December 31, 2023, whichever occurred first.

### Statistical Analysis

Baseline characteristics were summarized as mean ± standard deviation, median (interquartile range), or frequency (percentage), as appropriate. For the baseline renin analysis, Cox proportional hazards models were used to estimate adjusted hazard ratios (HRs) and 95% confidence intervals (CIs) for associations between baseline plasma renin activity and clinical outcomes. Models were adjusted for baseline age, sex, body mass index, serum creatinine, diabetes mellitus, heart failure, and antihypertensive medication classes and number. The continuous association between plasma renin activity and MACE was evaluated using restricted cubic splines with four knots placed at the 5th, 35th, 65th, and 95th percentiles of log-transformed renin values. Renin categories were informed by the observed spline association and defined as very low <0.3, low 0.3–1.0, moderately high >1.0–3.0 and high >3.0 ng/mL/h, with 0.3–1.0 ng/mL/h serving as the reference category. For the MRA analysis, time-dependent Cox models were used to estimate associations between MRA use and

MACE and its individual components within each baseline renin category. Models compared MRA-exposed with MRA-unexposed person-time and included MRA use as a time-varying exposure. Age and sex were included as baseline covariates, whereas body mass index, serum creatinine, diabetes mellitus, heart failure, and concomitant antihypertensive medication classes were included as time-varying covariates. The same model specification was applied in the sensitivity analysis described above. Missing baseline laboratory measurements and body mass index were handled using expectation-maximization single imputation before analysis. Restricted cubic spline modeling was performed using R software (version 4.5.3) with the rms package, and other analyses were performed using SAS version 9.4 (SAS Institute Inc., Cary, NC, USA). A two-sided P value <0.05 was considered statistically significant.

### Role of the funding source

The funders had no role in study design, data collection, data analysis, data interpretation, writing of the report, or the decision to submit the paper for publication.

## Results

### Study Population

A total of 19,448 patients who underwent plasma renin activity testing between January 2006 and December 2022 were initially identified. After excluding patients with prior adrenalectomy, baseline MRA use, baseline loop diuretic use, and missing aldosterone measurements, 16,600 patients were included in the final analysis (**Supplementary Figure 1**). Baseline characteristics are presented in **Table 1**.

### Association between Renin and Cardiovascular Outcomes

During a median follow-up of 3.8 years (interquartile range [IQR], 1.6 to 7.7 years), a total of 2564 MACE events occurred including 1687 all-cause deaths, 714 ischemic strokes, 658 hemorrhagic strokes and 123 acute myocardial infarctions. In RCS analysis, plasma renin demonstrated a U-shaped association with the risk of MACE with the lowest risk observed at approximately 1.17 ng/mL/h. The risk of MACE increased at plasma renin levels below 0.32 and above 3.36 ng/mL/h (**Figure 2A**).

**Figure 2.**
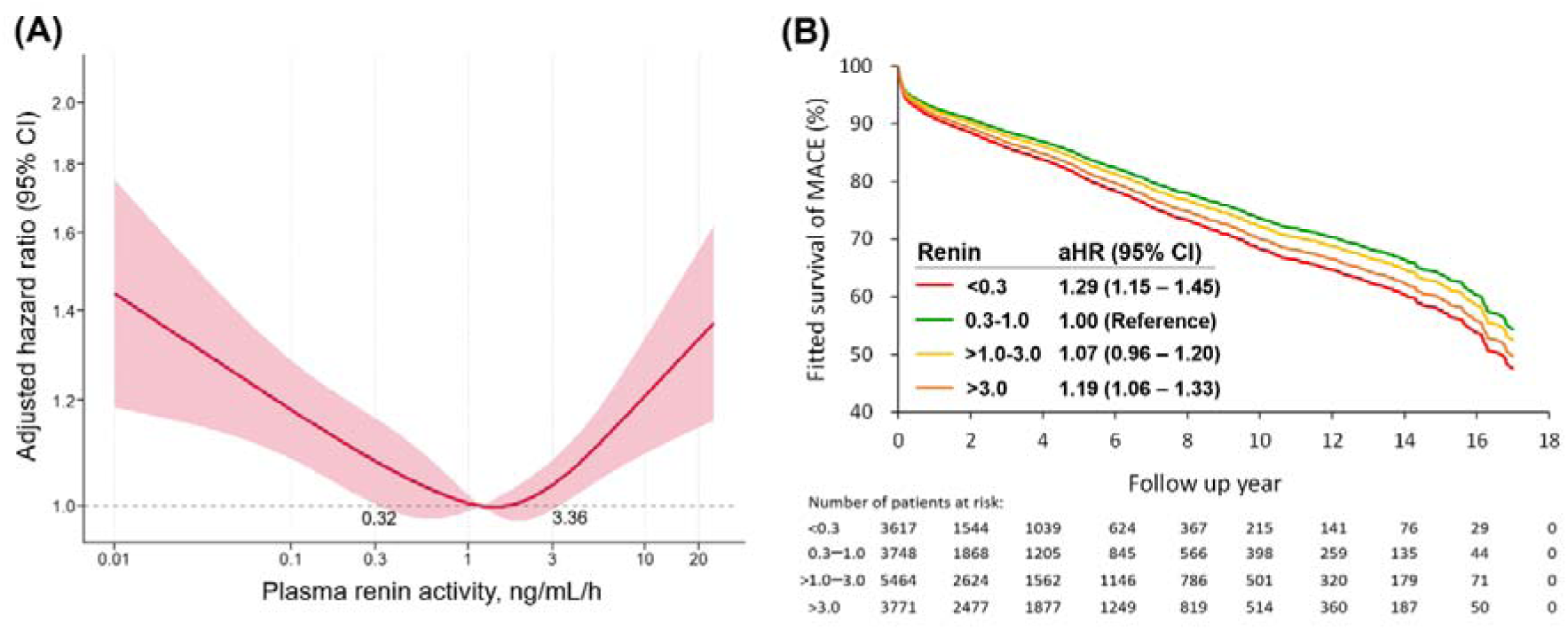
Association between plasma renin activity and risk of major adverse cardiovascular events. (A) Restricted cubic spline analysis demonstrating the association between baseline plasma renin activity and risk of major adverse cardiovascular events (MACE). Renin was log-transformed and modeled using Cox proportional hazards regression adjusted for age, sex, BMI, serum creatinine, diabetes, heart failure, antihypertensive medication classes and burden. The x-axis is presented in the original renin scale after back-transformation. Shaded areas indicate 95% confidence intervals. (B) Adjusted MACE-free survival curves and hazard ratios (95% CIs) across baseline plasma renin activity categories, with 0.3–1.0 ng/mL/h as the reference category. Curves and estimates were derived from Cox proportional hazards models adjusted for baseline age, sex, body mass index, serum creatinine, diabetes mellitus, heart failure, and antihypertensive medication classes and burden. Follow-up was censored at MRA initiation or adrenalectomy.

**Figure 3.**
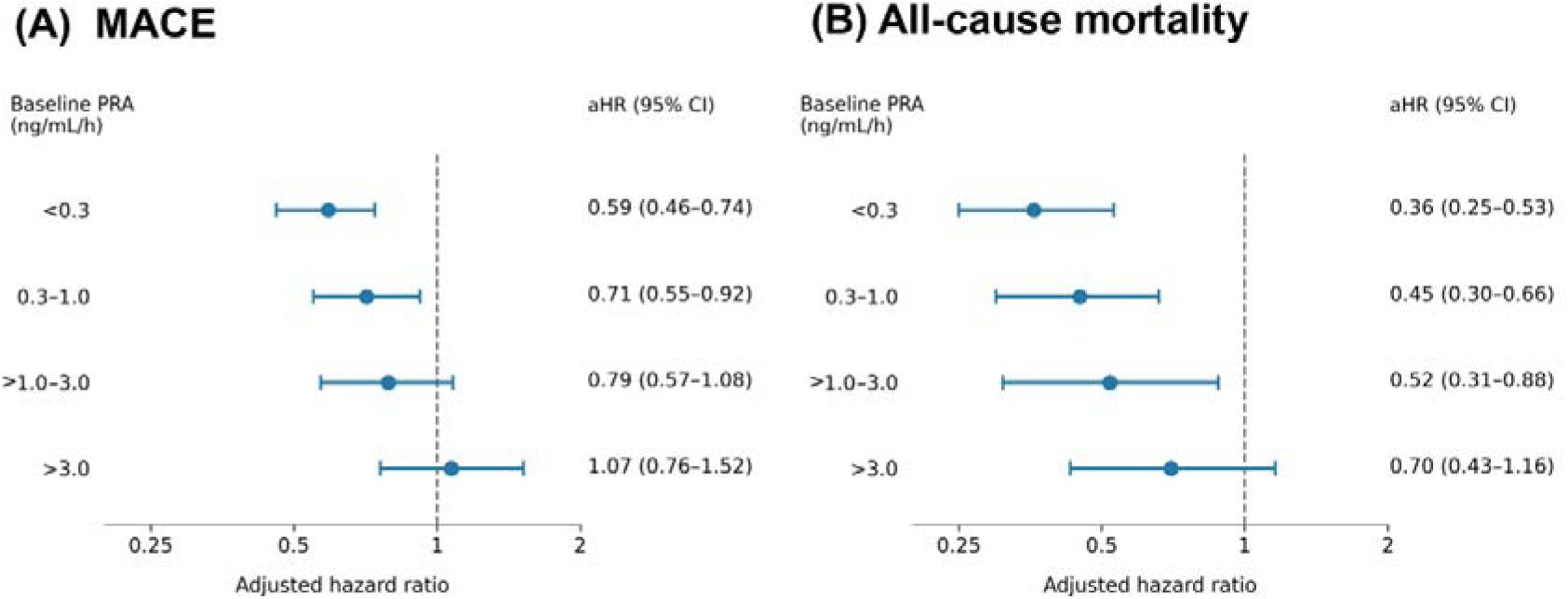
Associations of time-varying MRA use with MACE and all-cause mortality across baseline renin categories. Forest plots show adjusted hazard ratios for (A) major adverse cardiovascular events (MACE) and (B) all-cause mortality, comparing MRA use with non-use within each baseline plasma renin activity category. Estimates were obtained from time-dependent Cox models with MRA exposure modeled as a time-varying variable. Models were adjusted for baseline age and sex and time-varying body mass index, serum creatinine, diabetes mellitus, heart failure, and concomitant antihypertensive medication use, including renin-angiotensin system inhibitors, beta-blockers, alpha-blockers, vasodilators, thiazide diuretics, calcium channel blockers, clonidine, and minoxidil. Follow-up was censored at adrenalectomy.

Based on these observed thresholds, we categorized patients into four groups including very low renin (<0.3 ng/mL/h), low renin (0.3-1.0 ng/mL/h), moderately high renin (>1.0-3.0 ng/mL/h) and high renin (>3.0 ng/mL/h). Among them, 3,617 patients were classified as very low renin, 3,748 patients as low renin, 5,464 patients as moderately high renin, and 3,771 patients as high renin. Their clinical characteristics were listed in **Table 1**. Patients in the very low renin group were older and more likely to be female, whereas those in the high-renin group were younger and more likely to be male. Baseline cardiovascular risk factors and comorbidities were generally comparable across groups, with small standardized differences observed. Median plasma renin activity increased stepwise across renin categories, from 0.13 ng/mL/h (IQR, 0.08-0.20) in the very low renin group to 5.9 ng/mL/h (IQR, 4.4-7.1) in the high-renin group. Aldosterone levels showed a modest increase across renin categories, ranging from 22.9 ng/dL (IQR, 13.2-35.5) in the very low renin group to 33.1 ng/dL (IQR, 19.5-51.0) in the high-renin group, whereas aldosterone-to-renin ratios decreased progressively from 177.3 (IQR, 85.8-462.2) to 5.4 (IQR, 2.7-9.4).

The incidence of MACE across ascending renin categories was 5.87, 4.16, 3.92, and 3.61 events per 100 person-years, respectively (**Table 2**). After multivariable adjustment, both renin <0.3 ng/mL/h (HR, 1.29; 95% CI, 1.15–1.45) and renin >3.0 ng/mL/h (HR, 1.19; 95% CI, 1.06–1.33) were associated with higher MACE risk compared with the reference category of 0.3–1.0 ng/mL/h. No significant difference was observed for renin >1.0–3.0 ng/mL/h (HR, 1.07; 95% CI, 0.96–1.20) (**Table 2**, **Figure 2B**). For individual outcomes, both low and high baseline renin were associated with higher all-cause mortality compared with the 0.3–1.0 ng/mL/h category (very low renin: HR, 1.26; 95% CI, 1.09–1.45; high renin: HR, 1.28; 95% CI, 1.11–1.47). Very low renin was also associated with a higher risk of intracranial hemorrhage (HR, 1.48; 95% CI, 1.17–1.86). No statistically significant associations were observed for ischemic stroke or acute myocardial infarction (**Table 2**).

**Table 2.** Renin-stratified incidence and adjusted risk of major adverse cardiovascular events.

|  | Incidence (95% CI)‡ |  |  |  | Adjusted HR (95% CI)† |  |  |
|---|---|---|---|---|---|---|---|
| Outcome | Renin <0.3<br>(*n* = 3,617) | Renin 0.3–1.0<br>(*n* = 3,748) | Renin >1.0–3.0<br>(*n* = 5,464) | Renin >3.0<br>(*n* = 3,771) | <0.3 vs<br>0.3–1.0 | >1.0–3.0 vs<br>0.3–1.0 | >3.0 vs<br>0.3–1.0 |
| MACE | 5.87 (5.40–6.33) | 4.16 (3.82–4.51) | 3.92 (3.64–4.20) | 3.61 (3.33–3.89) | 1.29 (1.15–1.45)* | 1.07 (0.96–1.20) | 1.19 (1.06–1.33)* |
| Ischemic stroke | 1.60 (1.36–1.85) | 1.28 (1.09–1.47) | 0.99 (0.85–1.14) | 0.96 (0.82–1.10) | 1.13 (0.91–1.40) | 0.85 (0.69–1.05) | 0.96 (0.78–1.19) |
| Intracranial hemorrhage | 1.51 (1.28–1.74) | 0.92 (0.76–1.08) | 1.04 (0.89–1.18) | 0.89 (0.76–1.03) | 1.48 (1.17–1.86)* | 1.12 (0.90–1.40) | 1.22 (0.97–1.54) |
| Acute myocardial infarction | 0.21 (0.12–0.29) | 0.19 (0.12–0.26) | 0.20 (0.14–0.27) | 0.17 (0.11–0.23) | 1.06 (0.61–1.85) | 1.15 (0.70–1.87) | 0.99 (0.59–1.66) |
| All-cause death | 3.62 (3.27–3.97) | 2.61 (2.34–2.87) | 2.35 (2.14–2.57) | 2.32 (2.11–2.54) | 1.26 (1.09–1.45)* | 1.12 (0.98–1.29) | 1.28 (1.11–1.47)* |

Abbreviation: CI, confidence interval; HR, hazard ratio; MACE, major adverse cardiovascular events.
‡ Number of events per 100 person-year.
† Models were adjusted for age, sex, body mass index, serum creatinine, diabetes mellitus, heart failure, and baseline antihypertensive medication classes and antihypertensive medication burden at baseline.
\* Denotes *P* <0.05.

### Associations of MRA Use With Outcomes Across Baseline Renin Categories

In time-varying analyses, MRA use was associated with lower MACE risk in patients with very low baseline renin, <0.3 ng/mL/h (HR, 0.59; 95% CI, 0.46–0.74; P <0.001) and low baseline renin 0.3–1.0 ng/mL/h (HR, 0.71; 95% CI, 0.55–0.92; P = 0.010). No statistically significant association was observed in patients with moderately high renin >1.0–3.0 ng/mL/h (HR, 0.79; 95% CI, 0.57–1.08; P = 0.135) or high renin >3.0 ng/mL/h (HR, 1.07; 95% CI, 0.76–1.52; P = 0.697) (**Table 3**).

**Table 3.** Renin-stratified association of mineralocorticoid receptor antagonist use with major adverse cardiovascular events.

| MACE component/<br>renin | MRA-exposed intervals |  | MRA-unexposed intervals |  | Adjusted<br>HR (95% CI)† | *P* value |
|---|---|---|---|---|---|---|
|  | No. of<br>intervals | Incidence<br>(95% CI)‡ | No. of intervals | Incidence<br>(95% CI)‡ |  |  |
| MACE |  |  |  |  |  |  |
| <0.3 | 11408 | 2.93 (2.29–3.58) | 52753 | 5.69 (5.28–6.11) | 0.59 (0.46–0.74) | <0.001 |
| 0.3–1.0 | 10392 | 2.64 (2.00–3.28) | 63508 | 4.21 (3.88–4.54) | 0.71 (0.55–0.92) | 0.010 |
| >1.0–3.0 | 5287 | 3.25 (2.26–4.25) | 83403 | 4.04 (3.76–4.32) | 0.79 (0.57–1.08) | 0.135 |
| >3.0 | 3380 | 4.20 (2.79–5.62) | 79219 | 3.70 (3.43–3.98) | 1.07 (0.76–1.52) | 0.697 |
| Ischemic stroke |  |  |  |  |  |  |
| <0.3 | 11597 | 1.51 (1.06–1.97) | 53989 | 1.47 (1.26–1.68) | 0.95 (0.68–1.34) | 0.789 |
| 0.3–1.0 | 10537 | 1.10 (0.70–1.51) | 64395 | 1.23 (1.05–1.40) | 0.86 (0.57–1.28) | 0.447 |
| >1.0–3.0 | 5423 | 1.54 (0.87–2.22) | 85071 | 1.01 (0.88–1.15) | 1.30 (0.82–2.08) | 0.267 |
| >3.0 | 3477 | 1.92 (0.98–2.86) | 81035 | 0.97 (0.83–1.11) | 1.59 (0.95–2.68) | 0.078 |
| Intracranial hemorrhage |  |  |  |  |  |  |
| <0.3 | 11945 | 0.91 (0.56–1.26) | 54941 | 1.25 (1.06–1.44) | 0.72 (0.47–1.09) | 0.120 |
| 0.3–1.0 | 10755 | 0.73 (0.40–1.06) | 65625 | 0.80 (0.66–0.94) | 0.91 (0.56–1.48) | 0.695 |
| >1.0–3.0 | 5440 | 1.00 (0.46–1.54) | 85597 | 0.99 (0.85–1.13) | 0.77 (0.44–1.37) | 0.378 |
| >3.0 | 3621 | 0.92 (0.28–1.56) | 81836 | 0.89 (0.75–1.02) | 0.78 (0.38–1.59) | 0.488 |
| Acute myocardial infarction |  |  |  |  |  |  |
| <0.3 | 12128 | 0.14 (0.00–0.27) | 56298 | 0.20 (0.13–0.28) | 0.67 (0.23–1.97) | 0.471 |
| 0.3–1.0 | 10792 | 0.35 (0.12–0.57) | 66826 | 0.17 (0.11–0.24) | 2.06 (0.95–4.46) | 0.067 |
| >1.0–3.0 | 5619 | 0.15 (0.06–0.36) | 87146 | 0.21 (0.15–0.28) | 0.52 (0.12–2.18) | 0.369 |
| >3.0 | 3606 | 0.58 (0.07–1.08) | 83265 | 0.19 (0.13–0.25) | 2.43 (0.93–6.33) | 0.070 |
| All-cause death |  |  |  |  |  |  |
| <0.3 | 12209 | 0.96 (0.60–1.31) | 56556 | 3.85 (3.52–4.18) | 0.36 (0.25–0.53) | <0.001 |
| 0.3–1.0 | 10923 | 1.06 (0.67–1.46) | 67027 | 2.93 (2.66–3.19) | 0.45 (0.30–0.66) | <0.001 |
| >1.0–3.0 | 5638 | 1.11 (0.55–1.68) | 87586 | 2.56 (2.35–2.78) | 0.52 (0.31–0.88) | 0.015 |
| >3.0 | 3699 | 1.80 (0.92–2.68) | 83960 | 2.48 (2.26–2.70) | 0.70 (0.43–1.16) | 0.171 |

Intervals represent 3-month follow-up periods, classified according to MRA exposure. Counts refer to intervals rather than individual patients; each patient could contribute intervals to both exposure categories.
**Abbreviation:** CI, confidence interval; HR, hazard ratio; MACE, major adverse cardiovascular events; MRA, mineralocorticoid receptor antagonists.
‡ Number of events per 100 person-year.
† Models were adjusted for baseline age and sex and time-varying body mass index, serum creatinine, diabetes mellitus, heart failure, and concomitant antihypertensive medication use.

MRA use was also associated with lower all-cause mortality in patients with renin <0.3 ng/mL/h (HR, 0.36; 95% CI, 0.25–0.53; P <0.001), 0.3–1.0 ng/mL/h (HR, 0.45; 95% CI, 0.30–0.66; P <0.001), and >1.0–3.0 ng/mL/h (HR, 0.52; 95% CI, 0.31–0.88; P = 0.015). The association was not statistically significant in patients with renin >3.0 ng/mL/h (HR, 0.70; 95% CI, 0.43–1.16; P = 0.171). For intracranial hemorrhage, point estimates consistently favored a lower risk associated with MRA use across renin categories. However, none of these associations reached statistical significance. Associations with ischemic stroke and acute myocardial infarction were also not statistically significant within any renin category (**Table 3**). Findings were similar in sensitivity analyses without censoring at adrenalectomy. MRA use remained associated with lower MACE risk at baseline renin <0.3 ng/mL/h (HR, 0.60; 95% CI, 0.48–0.76) and 0.3–1.0 ng/mL/h (HR, 0.72; 95% CI, 0.56–0.93). Associations with lower all-cause mortality also persisted across renin categories ≤3.0 ng/mL/h (**Supplementary Table 2**).

## Discussion

In this large multicenter cohort of 16,600 patients with hypertension, baseline plasma renin activity demonstrated a U-shaped association with MACE, with the lowest estimated risk at approximately 1.17 ng/mL/h (remarkably similar to the threshold of 1.0 ng/mL/h identified in prior studies in people with primary aldosteronism^8,11^). In time-varying analyses, MRA use was associated with lower MACE risk in patients with very low or low renin, <0.3 and 0.3–1.0 ng/mL/h, respectively, whereas no statistically significant association was observed when renin was high. MRA use was also associated with lower all-cause mortality across renin categories ≤3.0 ng/mL/h. These findings suggest that renin may provide prognostic information and that MRA therapy may bestow its largest benefits in people with a low-renin phenotype of hypertension, as has been suggested in resistant hypertension^27,28^.

An important finding of this study is the marked heterogeneity in treatment response across renin-defined phenotypes. In patients with lower renin levels, initiation of MRAs was associated with a significant reduction in MACE risk. This observation supports the concept that a low renin identifies a hypertensive phenotype characterized by relative mineralocorticoid excess, even in the absence of a diagnosis of known PA. In this context, MRAs might not be viewed solely as add-on therapy for resistant hypertension, but rather as an early or upfront, mechanism-based treatment strategy for a substantial subset of patients. These findings further support the concept that low-renin hypertension is increasingly recognized as a heterogeneous condition encompassing a pathophysiologic spectrum of renin-independent aldosteronism and mineralocorticoid receptor overactivation^15,32–35^.

Clinical trials have demonstrated the preferential efficacy of mineralocorticoid receptor antagonists in lowering blood pressure in low-renin hypertension for over 50 years^4,36^. Although recent guidelines have proposed a more pragmatic diagnostic framework and a more liberal use of MRAs in PA, a substantial proportion of patients with low-renin hypertension do not meet conventional diagnostic criteria for PA^10^, even though recent studies indicate that they probably reside on the PA spectrum^37^. At the higher end of the renin spectrum (>3.0 ng/mL/h), however, MRA use was not significantly associated with MACE or all-cause mortality. This may suggest that a high renin state may be better treated with medications that interrupt the renin-angiotensin system (such as ACE inhibitors and/or angiotensin receptor blockers) rather than MRAs. Although the confidence intervals do not exclude potential benefit or harm, these results suggest that the favorable associations observed in patients with low renin may not extend uniformly across the renin spectrum. This pattern supports further investigation of baseline renin as a potential marker for identifying patients most likely to benefit from MRA therapy.

It is important to note that our results may be susceptible to confounding by indication. In clinical practice, MRAs may be prescribed to patients with resistant hypertension or heart failure, whose underlying cardiovascular risk may differ from that of patients not receiving MRAs. By modeling MRA exposure and relevant clinical covariates over time, we accounted for changes in treatment status, comorbidities, and concomitant antihypertensive medication use during follow-up. Nevertheless, this approach cannot fully account for differences in disease severity or the clinical factors influencing treatment decisions. The associations observed across renin categories therefore warrant further investigation rather than establishing differential treatment effects. While current guidelines recommend renin and aldosterone testing in all hypertensive patients primarily for the detection of PA^1,9,10,38^, our findings suggest a broader prognostic role of testing. Plasma renin may serve not only as a diagnostic tool but also as a practical biomarker for risk stratification and treatment guidance in hypertension.

Our findings build on the classical concept of renin profiling proposed by Laragh and colleagues, in which plasma renin activity helps distinguish volume-dependent from vasoconstriction-driven hypertension^13^. Previous studies have largely evaluated renin-guided treatment in relation to blood pressure responses^5,16^. In contrast, the present study contributes observational evidence concerning long-term clinical outcomes. It also relates to the evolving concept of a continuum of renin-independent aldosterone production rather than a strict dichotomy between PA and other forms of hypertension^18,35,39–42^. In this context, renin suppression may serve as a practical and clinically accessible marker to identify patients with hypertension who either have undiagnosed PA or those who could benefit from early initiation of aldosterone-targeted therapy, even in the absence of a formal PA diagnosis. This approach shifts the focus from diagnostic labeling toward therapeutic targeting of underlying biology to mitigate risk. However, our study did not directly compare a renin-guided treatment strategy with usual care. Further prospective studies are needed to confirm these observational associations.

Several limitations of this study should be acknowledged. First, treatment was not randomized, and residual confounding remains possible. Our time-dependent models incorporated changes in both MRA exposure and clinical covariates, an important methodological strength compared with analyses relying solely on baseline information. However, this approach cannot fully eliminate confounding by indication or bias arising when time-varying covariates are affected by prior treatment. Treatment discontinuation during clinical deterioration may also influence observed associations, particularly those involving mortality. Second, plasma renin measurements were obtained in real-world clinical settings without standardized medication washout or uniform sampling conditions. Although this may introduce variability and potential misclassification of renin phenotypes, it also reflects routine clinical practice and enhances the generalizability of our findings. Importantly, such non-differential variability would be expected to bias the results toward the null. In addition, current clinical guidelines support renin and aldosterone testing without strict medication withdrawal, which is consistent with our study design^10^. Third, the long study period and multicenter design also introduce potential measurement heterogeneity across sites and over time. Fourth, prescription-based exposure does not establish actual medication adherence or adequate mineralocorticoid receptor blockade. The present analyses did not evaluate whether dose titration or biochemical responses modified outcomes. Prior studies suggest that outcomes during MRA therapy may be related to the adequacy of treatment and post-treatment renin status ^8,11,43,44^. Our findings should therefore be interpreted as associations with recorded MRA use rather than the effects of optimized therapy. Fifth, while our findings support a renin-guided approach to antihypertensive therapy, causal inference cannot be established. In particular, the observed differential treatment effects across renin strata should be considered hypothesis-generating. Though prospective randomized trials have evaluated this strategy with respect to short-term blood pressure control^14,27,28,36^, long-term studies with clinical outcome measures are still needed.

In conclusion, renin may serve as a prognostic biomarker linking the underlying pathophysiology of hypertension, including unrecognized PA pathophysiology, to long-term clinical risk. In this large multicenter study, lower renin levels were associated with higher MACE risk, while MRA use in these individuals was associated with lower MACE risk.

These findings support a potential role for renin-guided medical therapy in general hypertension, where a low-renin phenotype may identify individuals who particularly benefit from an MRA rather than an agnostic approach to pharmacologic therapy.

## Funding

CHT was supported by Taiwan Society of Cardiology, the National Science and Technology Council, Taiwan grant 113-2314-B-002 -152 -MY2 and National Taiwan University Hospital grant 113-M0023, 114-M0030. AV was supported by National Institutes of Health awards R01DK115392, R01HL153004, R01HL155834.

## Data sharing

The individual-level data underlying this study are not publicly available because of patient confidentiality and institutional restrictions. Requests for data access can be directed to the corresponding author and will be considered subject to approval by National Taiwan University Hospital and applicable ethical and data protection requirements.

## Contributors

CHT, YHL and AV conceived and designed the study. CHT contributed to data acquisition and curation. CHT performed the statistical analyses. CHT, YHL and AV drafted the manuscript. All authors contributed to the interpretation of the findings and critically revised the manuscript for important intellectual content. CHT and YHL accessed and verified the underlying data. All authors approved the final manuscript and had final responsibility for the decision to submit for publication.

## Declaration of interests

AV reports consulting fees unrelated to the contents of this work from Corcept Therapeutics, Mineralys, HRA Pharma, Moderna, SideraBio Vertex, AstraZeneca, Adaptyx. JMB reports consulting fees unrelated to the contents of this work from Recordati Rare Diseases, Mineralys and AstraZeneca. All other coauthors have nothing to disclose.

## Acknowledgements

We thank the staff of the Eighth and Second Core Lab in the Department of Medical Research at National Taiwan University Hospital for technical support. We also thank all members of the TAIPAI Study Group (https://doi.org/10.6084/m9.figshare.21669929.v7) for help during the study. We would like to express our thanks to the staff of Department of Medical Research for providing clinical data from National Taiwan University Hospital-integrative Medical Data Center. The authors also thank Alfred Hsing-Fen Lin, Zoe Ya-Zhu Syu, and Winnie Su-Hui Wu, who served in Raising Statistics Consultant Inc., for their assistance with statistical analysis.

## Notes

### Author Declarations

The study protocol was approved by the Institutional Review Board of National Taiwan University Hospital (IRB number: 202504119RINB), and the requirement for informed consent was waived due to the retrospective nature of the study.

### Summary of Updates

We have updated the category names and adjusted the manuscript formatting for clarity and consistency.

## References

1. McEvoy JW, McCarthy CP, Bruno RM, Brouwers S, Canavan MD, Ceconi C, et al. 2024 ESC Guidelines for the management of elevated blood pressure and hypertension. Eur Heart J 2024;45:3912–4018. doi: 10.1093/eurheartj/ehae178

2. Kario K, Okura A, Hoshide S, Mogi M. The WHO Global report 2023 on hypertension warning the emerging hypertension burden in globe and its treatment strategy. Hypertens Res 2024;47:1099–1102. doi: 10.1038/s41440-024-01622-w

3. Buhler FR, Laragh JH, Baer L, Vaughan ED, Jr., Brunner HR. Propranolol inhibition of renin secretion. A specific approach to diagnosis and treatment of renin-dependent hypertensive diseases. N Engl J Med 1972;287:1209–1214. doi: 10.1056/NEJM197212142872401

4. Adlin EV, Marks AD, Channick BJ. Spironolactone and hydrochlorothiazide in essential hypertension. Blood pressure response and plasma renin activity. Arch Intern Med 1972;130:855–858.

5. Preston RA, Materson BJ, Reda DJ, Williams DW, Hamburger RJ, Cushman WC, et al. Age-race subgroup compared with renin profile as predictors of blood pressure response to antihypertensive therapy. Department of Veterans Affairs Cooperative Study Group on Antihypertensive Agents. JAMA 1998;280:1168–1172. doi: 10.1001/jama.280.13.1168

6. Viola A, Monticone S, Burrello J, Buffolo F, Lucchiari M, Rabbia F, et al. Renin and aldosterone measurements in the management of arterial hypertension. Horm Metab Res 2015;47:418–426. doi: 10.1055/s-0035-1548868

7. Monticone S, D’Ascenzo F, Moretti C, Williams TA, Veglio F, Gaita F, et al. Cardiovascular events and target organ damage in primary aldosteronism compared with essential hypertension: a systematic review and meta-analysis. Lancet Diabetes Endocrinol 2018;6:41–50. doi: 10.1016/S2213-8587(17)30319-4

8. Hundemer GL, Curhan GC, Yozamp N, Wang M, Vaidya A. Cardiometabolic outcomes and mortality in medically treated primary aldosteronism: a retrospective cohort study. Lancet Diabetes Endocrinol 2018;6:51–59. doi: 10.1016/S2213-8587(17)30367-4

9. Writing Committee M, Jones DW, Ferdinand KC, Taler SJ, Johnson HM, Shimbo D, et al. 2025 AHA/ACC/AANP/AAPA/ABC/ACCP/ACPM/AGS/AMA/ASPC/NMA/PCNA/SGIM Guideline for the Prevention, Detection, Evaluation and Management of High Blood Pressure in Adults: A Report of the American College of Cardiology/American Heart Association Joint Committee on Clinical Practice Guidelines. Hypertension 2025;82:e212–e316. doi: 10.1161/HYP.0000000000000249

10. Adler GK, Stowasser M, Correa RR, Khan N, Kline G, McGowan MJ, et al. Primary Aldosteronism: An Endocrine Society Clinical Practice Guideline. J Clin Endocrinol Metab 2025;110:2453–2495. doi: 10.1210/clinem/dgaf284

11. Hundemer GL, Curhan GC, Yozamp N, Wang M, Vaidya A. Incidence of Atrial Fibrillation and Mineralocorticoid Receptor Activity in Patients With Medically and Surgically Treated Primary Aldosteronism. JAMA Cardiol 2018;3:768–774. doi: 10.1001/jamacardio.2018.2003

12. Vaughan ED, Jr., Laragh JH, Gavras I, Buhler FR, Gavras H, Brunner HR, et al. Volume factor in low and normal renin essential hypertension. Treatment with either spironolactone or chlorthalidone. Am J Cardiol 1973;32:523–532. doi: 10.1016/s0002-9149(73)80044-x

13. Laragh JH, Letcher RL, Pickering TG. Renin profiling for diagnosis and treatment of hypertension. JAMA 1979;241:151–156.

14. Weinberger MH, White WB, Ruilope LM, MacDonald TM, Davidson RC, Roniker B, et al. Effects of eplerenone versus losartan in patients with low-renin hypertension. Am Heart J 2005;150:426–433. doi: 10.1016/j.ahj.2004.12.005

15. Shah SS, Fuller PJ, Young MJ, Yang J. Update on Low-Renin Hypertension: Current Understanding and Future Direction. Hypertension 2024;81:2038–2048. doi: 10.1161/HYPERTENSIONAHA.124.23385

16. Shah SS, Zhang J, Gwini SM, Young MJ, Fuller PJ, Yang J. Efficacy and safety of mineralocorticoid receptor antagonists for the treatment of low-renin hypertension: a systematic review and meta-analysis. J Hum Hypertens 2024;38:383–392. doi: 10.1038/s41371-023-00891-1

17. Brown JM, Siddiqui M, Calhoun DA, Carey RM, Hopkins PN, Williams GH, et al. The Unrecognized Prevalence of Primary Aldosteronism: A Cross-sectional Study. Ann Intern Med 2020;173:10–20. doi: 10.7326/M20-0065

18. Tsai CH, Brown JM, Parisien-La Salle S, Newman AJ, Wu VC, Lin YH, et al. ACE Inhibition to Distinguish Low-Renin Hypertension From Primary Aldosteronism. Hypertension 2025;82:1046–1055. doi: 10.1161/HYPERTENSIONAHA.125.24711

19. Brown JM, Robinson-Cohen C, Luque-Fernandez MA, Allison MA, Baudrand R, Ix JH, et al. The Spectrum of Subclinical Primary Aldosteronism and Incident Hypertension: A Cohort Study. Ann Intern Med 2017;167:630–641. doi: 10.7326/M17-0882

20. Inoue K, Goldwater D, Allison M, Seeman T, Kestenbaum BR, Watson KE. Serum Aldosterone Concentration, Blood Pressure, and Coronary Artery Calcium: The Multi-Ethnic Study of Atherosclerosis. Hypertension 2020;76:113–120. doi: 10.1161/HYPERTENSIONAHA.120.15006

21. Hu J, Shen H, Huo P, Yang J, Fuller PJ, Wang K, et al. Heightened Cardiovascular Risk in Hypertension Associated With Renin-Independent Aldosteronism Versus Renin-Dependent Aldosteronism: A Collaborative Study. J Am Heart Assoc 2021;10:e023082. doi: 10.1161/JAHA.121.023082

22. Hundemer GL, Agharazii M, Madore F, Vaidya A, Brown JM, Leung AA, et al. Subclinical Primary Aldosteronism and Cardiovascular Health: A Population-Based Cohort Study. Circulation 2024;149:124–134. doi: 10.1161/CIRCULATIONAHA.123.066389

23. Grim C, Winnacker J, Peters T, Gilbert G. Low renin, "normal" aldosterone and hypertension: circadian rhythm of renin, aldosterone, cortisol and growth hormone. J Clin Endocrinol Metab 1974;39:247–256. doi: 10.1210/jcem-39-2-247

24. Adlin EV, Marks AD, Channick BJ. The salivary sodium/potassium ratio in hypertension: relation to race and plasma renin activity. Clin Exp Hypertens A 1982;4:1869–1880. doi: 10.3109/10641968209061646

25. Mackenzie IS, Brown MJ. Molecular and clinical investigations in patients with low-renin hypertension. Clin Exp Nephrol 2009;13:1–8. doi: 10.1007/s10157-008-0071-4

26. Ferreira JP, Collier T, Clark AL, Mamas MA, Rocca HB, Heymans S, et al. Spironolactone effect on the blood pressure of patients at risk of developing heart failure: an analysis from the HOMAGE trial. Eur Heart J Cardiovasc Pharmacother 2022;8:149–156. doi: 10.1093/ehjcvp/pvab031

27. Williams B, MacDonald TM, Morant S, Webb DJ, Sever P, McInnes G, et al. Spironolactone versus placebo, bisoprolol, and doxazosin to determine the optimal treatment for drug-resistant hypertension (PATHWAY-2): a randomised, double-blind, crossover trial. Lancet 2015;386:2059–2068. doi: 10.1016/S0140-6736(15)00257-3

28. Williams B, MacDonald TM, Morant SV, Webb DJ, Sever P, McInnes GT, et al. Endocrine and haemodynamic changes in resistant hypertension, and blood pressure responses to spironolactone or amiloride: the PATHWAY-2 mechanisms substudies. Lancet Diabetes Endocrinol 2018;6:464–475. doi: 10.1016/S2213-8587(18)30071-8

29. Cheng CL, Lee CH, Chen PS, Li YH, Lin SJ, Yang YH. Validation of acute myocardial infarction cases in the national health insurance research database in taiwan. J Epidemiol 2014;24:500–507. doi: 10.2188/jea.je20140076

30. Hsieh CY, Chen CH, Li CY, Lai ML. Validating the diagnosis of acute ischemic stroke in a National Health Insurance claims database. J Formos Med Assoc 2015;114:254–259. doi: 10.1016/j.jfma.2013.09.009

31. Hung LC, Sung SF, Hsieh CY, Hu YH, Lin HJ, Chen YW, et al. Validation of a novel claims-based stroke severity index in patients with intracerebral hemorrhage. J Epidemiol 2017;27:24–29. doi: 10.1016/j.je.2016.08.003

32. Baudrand R, Vaidya A. The Low-Renin Hypertension Phenotype: Genetics and the Role of the Mineralocorticoid Receptor. Int J Mol Sci 2018;19. doi: 10.3390/ijms19020546

33. Monticone S, Losano I, Tetti M, Buffolo F, Veglio F, Mulatero P. Diagnostic approach to low-renin hypertension. Clin Endocrinol (Oxf) 2018;89:385–396. doi: 10.1111/cen.13741

34. Vaidya A, Mulatero P, Baudrand R, Adler GK. The Expanding Spectrum of Primary Aldosteronism: Implications for Diagnosis, Pathogenesis, and Treatment. Endocr Rev 2018;39:1057–1088. doi: 10.1210/er.2018-00139

35. Vaidya A, Brown JM. Redefining Primary Aldosteronism. J Am Coll Cardiol 2026. doi: 10.1016/j.jacc.2026.02.5090

36. Carey RM, Douglas JG, Schweikert JR, Liddle GW. The syndrome of essential hypertension and suppressed plasma renin activity. Normalization of blood pressure with spironolactone. Arch Intern Med 1972;130:849–854.

37. Voorhies KR, Bi C, Adeyemo MA, Dixit SB, Vaidya A, McPhaul MJ, et al. Evaluation of Current Screening Thresholds for the Detection of Primary Aldosteronism. Hypertension 2026. doi: 10.1161/HYPERTENSIONAHA.126.27634

38. Naruse M, Katabami T, Shibata H, Sone M, Takahashi K, Tanabe A, et al. Japan Endocrine Society clinical practice guideline for the diagnosis and management of primary aldosteronism 2021. Endocr J 2022;69:327–359. doi: 10.1507/endocrj.EJ21-0508

39. Parksook WW, Brown JM, Omata K, Tezuka Y, Ono Y, Satoh F, et al. The Spectrum of Dysregulated Aldosterone Production: An International Human Physiology Study. J Clin Endocrinol Metab 2024;109:2220–2232. doi: 10.1210/clinem/dgae145

40. Vaidya A, Brown JM, Carey RM, Siddiqui M, Williams GH. The Unrecognized Prevalence of Primary Aldosteronism. Ann Intern Med 2020;173:683. doi: 10.7326/L20-1097

41. Ljungberg C, Carlsson B, Shojaiyan P, Franzen S, Bech JN, Chin KL, et al. Renin-Aldosterone Profiles and Cardiorenal Outcomes in Hypertension: A Nationwide Cohort Study. J Am Coll Cardiol 2026. doi: 10.1016/j.jacc.2026.01.056

42. Lassen MCH, Ostrominski JW, Claggett BL, Wijkman MO, Lee J, Van’t Hof JR, et al. Spectrum of Primary Aldosteronism and Risk of Cardiovascular Outcomes: The Atherosclerosis Risk in Communities Study. JAMA Cardiol 2026;11:341–351. doi: 10.1001/jamacardio.2026.0068

43. Chen ZW, Pan CT, Liao CW, Tsai CH, Chang YY, Chang CC, et al. Implication of MR Activity in Posttreatment Arterial Stiffness Reversal in Patients With Primary Aldosteronism. J Clin Endocrinol Metab 2023;108:624–632. doi: 10.1210/clinem/dgac649

44. Katsuragawa S, Le MV, Fuller PJ, Yang J. Post-treatment renin status and cardiovascular, renal, and mortality outcomes in medically treated primary aldosteronism: a systematic review and meta-analysis. Lancet Diabetes Endocrinol 2025;13:1041–1053. doi: 10.1016/S2213-8587(25)00263-3

