## Supplementary Figure 1 and Table 1-2 for "Renin, Major Adverse Cardiovascular Outcomes, and the Use of Mineralocorticoid Receptor Antagonists in Hypertension"

**Supplementary Figure 1 Study flowchart**

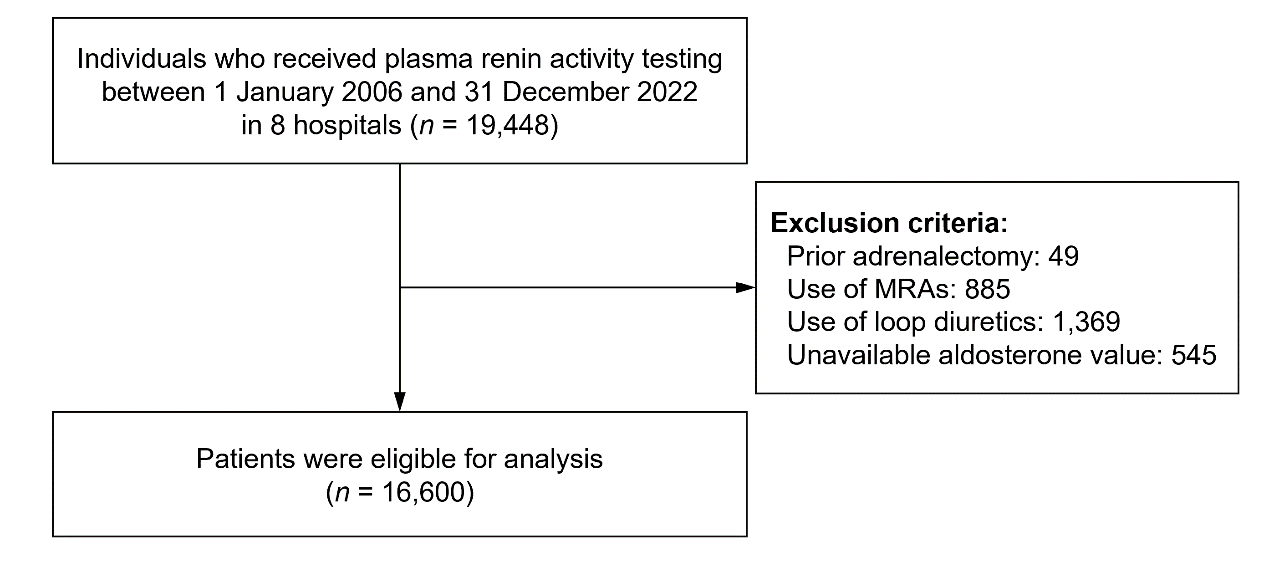
Flowchart of patient selection from the multicenter electronic medical record database. A total of 16,600 patients were included in the analysis.

**Supplemental Table 1.** ICD diagnostic codes used in the study

| Disease | ICD-9 | ICD-10 |
| --- | --- | --- |
| Hypertension | 401.x-405.x | I10-I15 |
| Diabetes mellitus | 250.x | E08-E13 |
| Hyperlipidemia | 272.0-272.4 | E77, E780, E781, E782, E783, E784, E785, E786, E881, E753, E755, E882, E756, E789, E7521, E7522, E7524, E7130, E7879, E7881, E7889, E8889, E7870 |
| Atrial fibrillation | 427.31 | I480-I482, I4891 |
| Obstructive sleep apnea | 327.23, 780.51, 780.53, 780.57 | G47.30, G47.31, G47.33, G47.37, G47.39 |
| Chronic obstructive pulmonary disease | 491.x, 492.x, 496 | J41.x, J42, J43.x, J44.x |
| Chronic kidney disease | 580.x–589.x, 403.x–404.x, 016.0x, 095.4x, 236.9x, 250.4x, 274.1x, 442.1x, 447.3x, 440.1x, 572.4x, 642.1x, 646.2x, 753.1x, 283.11, 403.01, 404.02, 446.21 | A1811, D593, E102, E112, E132, I12, I13, K767, M103, M310, N00, N01, N02, N03, N04, N05, N06, N07, N08, N14, N150, N158, N159, N16, N171, N172, N18, N19, N200, N25, N261, N269, N27, Q61 |
| Heart failure | 428.x | I50 |
| Acute myocardial infarction | 410.x | I21-I22 |
| Coronary artery disease | 410.x–414.x | I20-I24 |
| Intracranial hemorrhage | 430.x, 431.x, 432.x | I60-I62 |
| Ischemic stroke | 433.x–437.x | G450, G451, G452, G454, G458, G459, G460, G461, G462, G463, G464, G465, G466, G467, G468, I6300, I63011, I63012, I63019, I6302, I6303, I6309, I6310, I63111, I63112, I63119, I6312, I6313, I6319, I6320, I63211, I63212, I63219, I6322, I6323, I6329, I6330, I6331, I63311, I63312, I63319, I63321, I63322, I63329, I6333, I63331, I63332, I63339, I6334, I63341, I63342, I63349, I6339, I6340, I6341, I63411, I63412, I63419, I6342, I63421, I63422, I63429, I63431, I63432, I63439, I63441, I63442, I63449, I6349, I6350, I63511, I63512, I63519, I63521, I63522, I63529, I63531, I63532, I63539, I63541, I63542, I63549, I6359, I6359, I636, I638, I639, I650, I651, I6521, I6522, I6523, I6529, I658, I659, I66, I670, I671, I672, I674, I675, I676, I677, I6781, I6782, I6789, I679, I680, I682, I688 |

Abbreviation: ICD, International Classification of Diseases.

**Supplemental Table 2.** Renin-stratified association of mineralocorticoid receptor antagonist use with major adverse cardiovascular events, without censoring at the day of adrenalectomy

|  | MRA-exposed intervals | | MRA-unexposed intervals | | Adjusted  HR (95% CI)† | *P* value |
| --- | --- | --- | --- | --- | --- | --- |
| MACE component / renin | No. of intervals | Incidence  (95% CI)‡ | No. of intervals | Incidence  (95% CI)‡ |  |  |
| MACE |  |  |  |  |  |  |
| <0.3 | 11827 | 2.85 (2.23–3.47) | 60609 | 5.14 (4.77–5.51) | 0.60 (0.48–0.76) | <0.001 |
| 0.3–1.0 | 10736 | 2.59 (1.97–3.21) | 68037 | 4.06 (3.75–4.37) | 0.72 (0.56–0.93) | 0.012 |
| >1.0–3.0 | 5352 | 3.28 (2.28–4.27) | 86331 | 3.98 (3.71–4.26) | 0.80 (0.58–1.09) | 0.162 |
| >3.0 | 3573 | 4.19 (2.82–5.56) | 82125 | 3.61 (3.35–3.88) | 1.08 (0.77–1.51) | 0.673 |
| Ischemic stroke |  |  |  |  |  |  |
| <0.3 | 12027 | 1.49 (1.04–1.93) | 61942 | 1.32 (1.13–1.50) | 0.99 (0.71–1.40) | 0.997 |
| 0.3–1.0 | 10881 | 1.07 (0.67–1.46) | 68978 | 1.17 (1.01–1.34) | 0.85 (0.57–1.27) | 0.434 |
| >1.0–3.0 | 5488 | 1.52 (0.85–2.19) | 88010 | 0.99 (0.85–1.12) | 1.30 (0.82–2.08) | 0.266 |
| >3.0 | 3670 | 1.81 (0.92–2.70) | 83957 | 0.96 (0.82–1.09) | 1.49 (0.89–2.51) | 0.129 |
| Intracranial hemorrhage |  |  |  |  |  |  |
| <0.3 | 12385 | 0.87 (0.54–1.21) | 62989 | 1.13 (0.96–1.30) | 0.73 (0.48–1.11) | 0.142 |
| 0.3–1.0 | 11100 | 0.71 (0.39–1.03) | 70257 | 0.77 (0.64–0.90) | 0.89 (0.54–1.45) | 0.630 |
| >1.0–3.0 | 5505 | 0.98 (0.45–1.52) | 88620 | 0.96 (0.83–1.09) | 0.77 (0.44–1.37) | 0.375 |
| >3.0 | 3814 | 0.87 (0.27–1.47) | 84830 | 0.86 (0.74–0.99) | 0.76 (0.37–1.56) | 0.454 |
| Acute myocardial infarction |  |  |  |  |  |  |
| <0.3 | 12577 | 0.13 (0.00–0.26) | 64555 | 0.19 (0.12–0.26) | 0.62 (0.21–1.82) | 0.387 |
| 0.3–1.0 | 11137 | 0.33 (0.12–0.55) | 71558 | 0.16 (0.10–0.22) | 2.05 (0.95–4.43) | 0.067 |
| >1.0–3.0 | 5684 | 0.15 (0.06–0.35) | 90198 | 0.20 (0.14–0.27) | 0.52 (0.12–2.18) | 0.371 |
| >3.0 | 3809 | 0.54 (0.07–1.02) | 86350 | 0.18 (0.12–0.24) | 2.32 (0.89–6.04) | 0.085 |
| All-cause death |  |  |  |  |  |  |
| <0.3 | 12660 | 0.92 (0.58–1.26) | 64823 | 3.49 (3.20–3.78) | 0.37 (0.25–0.54) | <0.001 |
| 0.3–1.0 | 11268 | 1.06 (0.68–1.45) | 71764 | 2.83 (2.58–3.09) | 0.46 (0.32–0.67) | <0.001 |
| >1.0–3.0 | 5703 | 1.17 (0.60–1.74) | 90636 | 2.55 (2.34–2.76) | 0.55 (0.34–0.91) | 0.021 |
| >3.0 | 3902 | 1.91 (1.03–2.79) | 87058 | 2.42 (2.20–2.63) | 0.76 (0.47–1.23) | 0.261 |

Intervals represent 3-month follow-up periods, classified according to MRA exposure. Counts refer to intervals rather than individual patients; each patient could contribute intervals to both exposure categories.

**Abbreviation:** CI, confidence interval; HR, hazard ratio; MACE, major adverse cardiovascular events; MRA, mineralocorticoid receptor antagonists.

‡ Number of events per 100 person-year.

† Models were adjusted for baseline age and sex and time-varying body mass index, serum creatinine, diabetes mellitus, heart failure, and concomitant antihypertensive medication use.
